# Effects of PM_2.5_ from coal mine fire smoke on long-term incidence of major adverse cardiovascular events (MACE)

**DOI:** 10.64898/2026.01.28.26345097

**Authors:** Thara Govindaraju, Tyler J Lane, Matthew Carroll, Catherine L Smith, David Brown, David Poland, Jillian F Ikin, Alice J Owen, Thomas Wardill, Emily Nehme, Dion Stub, Michael J Abramson, Karen Walker-Bone, Tracy A McCaffrey, Caroline X Gao

## Abstract

**Background:** While coal mine fire smoke has been linked to short-term increases in cardiovascular events, there is little evidence on long-term risks. We investigated longer-term risk of major adverse cardiovascular events (MACE) following the 2014 Hazelwood coal mine fire in regional Victoria, Australia.

**Methods:** In this cohort study, combined administrative data on ambulance attendances, emergency department presentations, hospital admissions, and mortality from March 2014 to June 2022, with survey data from 2016/17. Time-location diaries for the mine-fire period were combined with modelled fire-related particulate matter ≤2.5µm in diameter (PM_2.5_) to estimate individual exposures. We analysed the association between PM_2.5_ exposure and time to MACE using a recurrent event survival analysis, adjusting for key confounders. Outcomes were examined over 8 years of follow-up and stratified by time.

**Results:** *N* = 2,725 cohort members agreed to linking their survey responses to administrative data. There was no detectable effect of fire-related PM_2.5_ exposure on overall risk of MACE during 8-year follow-up. However, there was weak evidence suggesting increase in MACE risk in the first 3 years post-fire, with hazard ratios ranging from 1.05-1.18 per 10µg/m^3^ of daily average PM_2.5_ exposure. Nearly all analyses of cardiovascular death detected an increased risk across the entire follow-up period, with hazard ratios ranging from 1.19-1.25 per 10µg/m^3^.

**Conclusions:** We found smoke exposure predicted an increase in cardiovascular health service use in the three years after the mine fire. There was additional evidence that the mine fire increased risk of cardiovascular death over the entire 8-year follow-up. This suggests that cardiovascular screening should be a routine component of planning recovery after landscape fires.

## Introduction

Landscape fires are a growing global threat. In Australia alone, the annual area burned by fire across Australia’s forests has been increasing by about 48,000 ha per year over the last three decades.^1^ Annually, more than 1.5 million deaths each year are attributed to air pollution from landscape fires.^2^ Climate change will exacerbate the frequency and intensity of these fires, resulting in excess morbidity and mortality globally.^3^

One of the most harmful air pollutants from landscape fires is fine particulate matter ≤2.5 micrometres in diameter, commonly referred to as PM_2.5_.^4^ Due to its small size, PM_2.5_ can penetrate deep into the airways where it then enters the blood stream and can be disseminated throughout the body. There is a substantial body of evidence that PM_2.5_ causes oxidative stress, inflammation, and stimulation of the autonomic nervous system leading to endothelial dysfunction, increasing the risk of cardiovascular diseases such as myocardial infarction, arrhythmia, heart failure, stroke, and cardiovascular-related deaths.^5–7^ There is also evidence that PM_2.5_ is more harmful when originating from landscape fires than sources such as traffic or industry, as it contains smaller and more toxic particles.^8–11^

### The Hazelwood coal mine fire and its effects on cardiovascular health

On February 9, 2014, the open cut brown coal mine adjacent to the town of Morwell in the Latrobe Valley, regional Victoria, caught fire, covering the nearby town of Morwell in plumes of smoke and ash over a six-week period.^12^ The Victorian State Government commissioned a long-term investigation into the effects of smoke exposure on the community, the Hazelwood Health Study.^13^

Investigations of mine fire impacts on cardiovascular health have produced inconsistent findings. For example, studies of effects on cardiovascular health services covering medications, hospital admissions, and ambulance attendances, have been mixed, indicating both an increase in use and no effect.^14–19^ However, health service use is an imperfect indicator of individual health as it is reliant upon willingness and self-efficacy, health literacy and the perceived severity of symptoms, as well as socioeconomic factors and service availability.^20^ Communities affected by the mine fire were characterised by already high service use^15^ as well as some of the highest levels of socioeconomic deprivation in Australia^21^ that only worsened with the closure of the coal mine and power plant in 2017.^22^

Previous assessments of cardiovascular disease risk markers in the Hazelwood Health Study failed to show any association with fire-related PM_2.5_ exposure.^23^ However, PM_2.5_ has been associated with an increased risk of cardiovascular death, with higher risk from PM_2.5_ originating from landscape fires.^11^ In the Hazelwood Health Study, we detected an increase in cardiovascular deaths in the 6 months following the fire in areas with the most smoke exposure,^24^ while the risk of cardiovascular death among individuals with the most smoke exposure remained elevated for the following 9 years.^25^ Unlike health service use, where there is some discretion (i.e., some people may not seek care for reasons unrelated to their health), death is nearly always recorded and is therefore a more robust outcome. The observed increase in cardiovascular deaths might be the result in reduced detection of severe non-fatal cardiovascular outcomes, and presents a challenge for research as those who were most vulnerable might not survive long enough to take part in surveys.

### This analysis

In this analysis, we focused on major adverse cardiovascular events (MACE), one of the most common primary endpoints in cardiovascular research.^26^ While not consistently defined,^26^ MACE can be thought of as the group of cardiovascular conditions that are most serious. The most commonly-used definitions for MACE include cardiovascular death, stroke (cerebrovascular accidents), and acute myocardial infarction; other, less commonly-used conditions include ischaemic heart disease and heart failure^26^ To the best of our knowledge, MACE have never been studied in the context of a medium-duration extreme exposure to coal mine fire-related PM_2.5_.^27^ Furthermore, MACE are serious and costly health events.^28^ It is important to know whether the treatment burden falls more heavily on one health service than another. To guide our approach, we addressed the following research questions: Did PM_2.5_ exposure from the Hazelwood coal mine fire increase the risk of MACE in the 8.5 years post-fire? And did this vary by health service?

## Methods

### Data sources

This study relied on data from the Hazelwood Health Study Adult Survey, which has been described previously.^29,30^ In summary, we used the Victorian Electoral Roll to identify and recruit 4056 adult residents of areas near the Hazelwood coal mine fire into a cohort known as the Adult Survey. This included 3096 residents of Morwell, the primary exposure site, and 960 residents of Sale, an unexposed town around 60km to the northeast. Of these, 2725 (2115 from Morwell, 610 from Sale) agreed to have their data linked to a range of administrative health datasets.

For this analysis, we linked Adult Survey participant responses^29^ to the Victorian Admitted Episodes Dataset (VAED) for hospital admissions,^31^ the Victorian Emergency Minimum Dataset (VEMD) for emergency department (ED) presentations,^32^ Ambulance Victoria’s electronic patient care records for ambulance attendances,^33^ as well as to the National Death Index (NDI) for deaths.^34^ All four data sources were linked to Adult Survey participants covering the period from January 1, 2009, to June 30, 2022, with the follow-up period defined as the interval between March 31, 2014 (end of the mine fire) and June 30, 2022.

### Exposure to mine fire-related PM_2.5_

To assess participant levels of exposure to fire-related PM_2.5_, we combined time-location diaries from the Adult Survey,^29^ which placed participants at 12-hour intervals during the mine fire, with air pollution data, generated from emissions and chemical transport models that factored in existing air monitoring data, local weather conditions, and coal combustion.^30,35^ The modelled air pollution data estimated spatial and temporal distributions of PM_2.5_ from the mine fire across Victoria, with resolution as precise as 100m^2^ in the areas surrounding the mine.^35^ The combination of the time-location diary and modelled air pollution data provided individual-level exposure estimates.

### Outcomes

The primary outcome was the time to MACE. For hospital admissions and ED presentations, MACE were identified through diagnoses of unstable angina, heart failure, cardiovascular death, acute myocardial infarction, stroke (not specified as haemorrhagic or infarction), cardiac arrest, cardiogenic shock, and ventricular fibrillation, based on the International Statistical Classification of Diseases and Related Health Problems, Tenth Revision, Australian Modification (ICD-10-AM). Comparable conditions were identified from ambulance attendances with reference to the final primary diagnosis (main condition identified by paramedics at the conclusion of their assessment and treatment of a patient). In addition, we included non-urgent hospital admissions for elective coronary revascularisation via percutaneous coronary intervention or bypass grafting, as defined by the Australian Classification of Health Interventions (ACHI).^36^ Finally, ICD-10-AM diagnoses for all cardiovascular conditions as the underlying cause of death in National Death Index records were coded as MACE. A complete list of all the diagnoses and procedure codes considered for the analysis is provided in Table S1.

As MACE has been defined in numerous ways across the academic literature,^26^ we developed several variations to test robustness of effects. The first variation included using either primary diagnoses (conditions that coexisted at the time of admission or developed subsequently and affected patient care, which also included the principal diagnosis) or principal diagnosis (the main condition causing need for health care) to identify MACE in hospital and ED records. The second involved capping MACE within a rolling 28-day period (e.g., if there was a MACE on day 1 and another on day 27, the second event would be combined with the first MACE to create one record). The aim of this second approach was to minimise double, triple, or quadruple counting across services, such as someone being transported via ambulance, presenting at ED, being admitted to hospital, and then dying, which could be recorded as four separate events. The third variation accounted for a lag between death notification and recording cause of death in the NDI, which affected the last 6 months of data. Using sociodemographic, behavioural, and health data from across the linked datasets, we predicted likely cardiovascular-related deaths a nested cross-validated (outer loop: 5-fold and inner loop: 10-fold) XGBoost algorithm.^37^ Lastly, to make the findings more comparable to the existing evidence base that has typically excluded non-emergency procedures,^26^ we conducted sensitivity analyses that limited MACE to urgent conditions, i.e., those not primarily intended as treatments or preventive measures.

### Confounders

We accounted for a range of potential confounders, all self-reported at the time of the Adult Survey, including; demographics (age, sex, marital status); pre-fire doctor-diagnosed health conditions (high blood pressure, high cholesterol, angina, other cardiovascular conditions, mental health, diabetes, asthma/chronic obstructive pulmonary disease[COPD]); smoking status (current, former, never); socioeconomic status based on the Index of Relative Socioeconomic Advantage and Disadvantage (IRSAD) score from the 2016 Census for the participant’s residential area;^21^ educational attainment (secondary to year 11–12, trade certification/university/tertiary education); and occupational exposures (working at least 6 months in a job with exposure to dust, fumes, gas, vapor or mist, separated into those related to coal mines or power stations and other industries). We also included the number of MACE events extracted from the linked data prior to the mine fire as a confounder.

### Analysis

Descriptive statistics characterised the sample by fire-related PM_2.5_ exposure levels (i.e., low, medium, and high exposure defined by tertiles). Continuous variables are presented as means and standard deviations, and categorical/dichotomous variables as counts and percentages.

We applied a recurrent event survival model, the Prentice, Williams and Peterson counting process (PWP-CP)^38^ method. This is a counting process-based extension of the Cox model that stratifies events (in this case, MACE) by their number, enabling event-specific baseline hazards within each stratum. As the proportional hazards assumption was violated, we estimated the time-stratified effects of exposure to the mine fire (first 3 years or ≥3 years). Exposure effects are presented as hazard ratios (HR) and 95% confidence intervals (CI) associated with every 10μg/m^3^ increase in PM_2.5_. All events were censored at either the date of death or the end of the follow-up period.

Cardiovascular-related ambulance, ED, hospital, and death data were combined to analyse as a single outcome (composite MACE) and were also analysed separately. We also analysed each approach to the MACE definition separately (based on primary versus principal diagnosis; capping events within a rolling 28 days; with predicted cardiovascular-related mortality). Analysis of MACE excluding non-emergency hospital procedures served as a sensitivity analysis i.e., restricted to ambulance, ED, death and emergency hospitalisations. Missing data were addressed using Multivariate Imputation by Chained Equations (MICE), producing 20 imputed datasets that were analysed separately and had their results pooled using Rubin’s rules.^39^ All analyses were conducted using R in RStudio.^40,41^

### Ethics

The study received approval from the Monash University Human Research Ethics committee under approval numbers CF15/872–2015000389 and 25680. All participants gave written informed consent for participation in the Adult Survey and for data linkage.

## Results

Table 1 describes the key characteristics of the cohort. Detailed cohort characteristics have been published elsewhere.^29,42^ There were several differences by exposure group, notably by educational attainment, marital status, and residential socioeconomic area (IRSAD score).

**Table 1.**
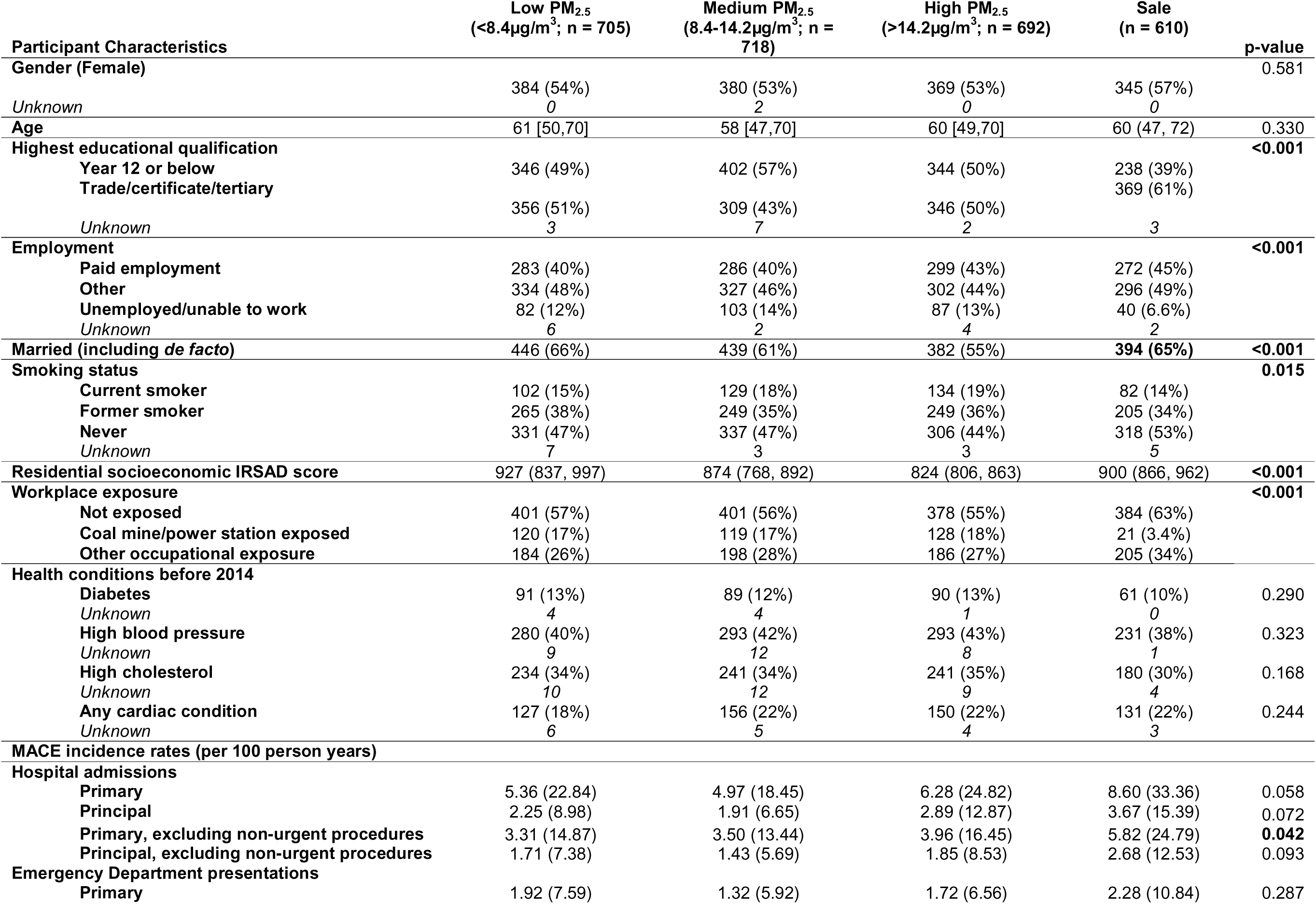
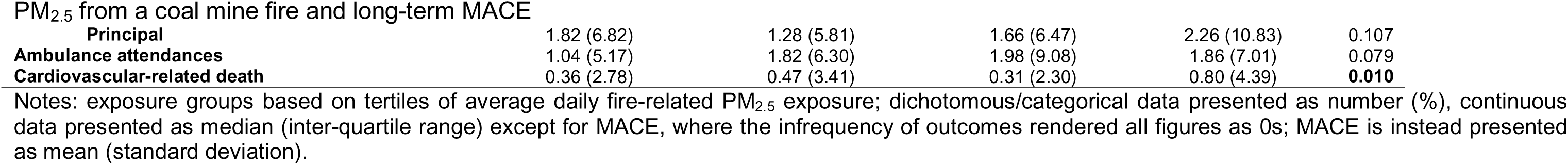
Descriptive statistics, grouped by fire-related related PM_2.5_ exposure.

Results of the recurrent events survival models are illustrated in Figure 1, with underlying data detailed in Tables S2 and S3. The results were consistent across different analyses, although the magnitude and statistical significance varied. In the first 3 years post-fire, there was weak evidence that composite MACE (across all services and including death) increased in association with a 10µg/m^3^ increase in mine fire-related PM_2.5_ (HR range: 1.05-1.12), which attenuated after 3 years. There was no evidence for an overall effect, i.e., across the entire study period. Sensitivity analyses excluding non-emergency hospital admissions produced similar results, though with a clearer effect on composite MACE in the first 3 years post-fire (HR range: 1.07-1.18); see Figure S1.

**Figure 1.**
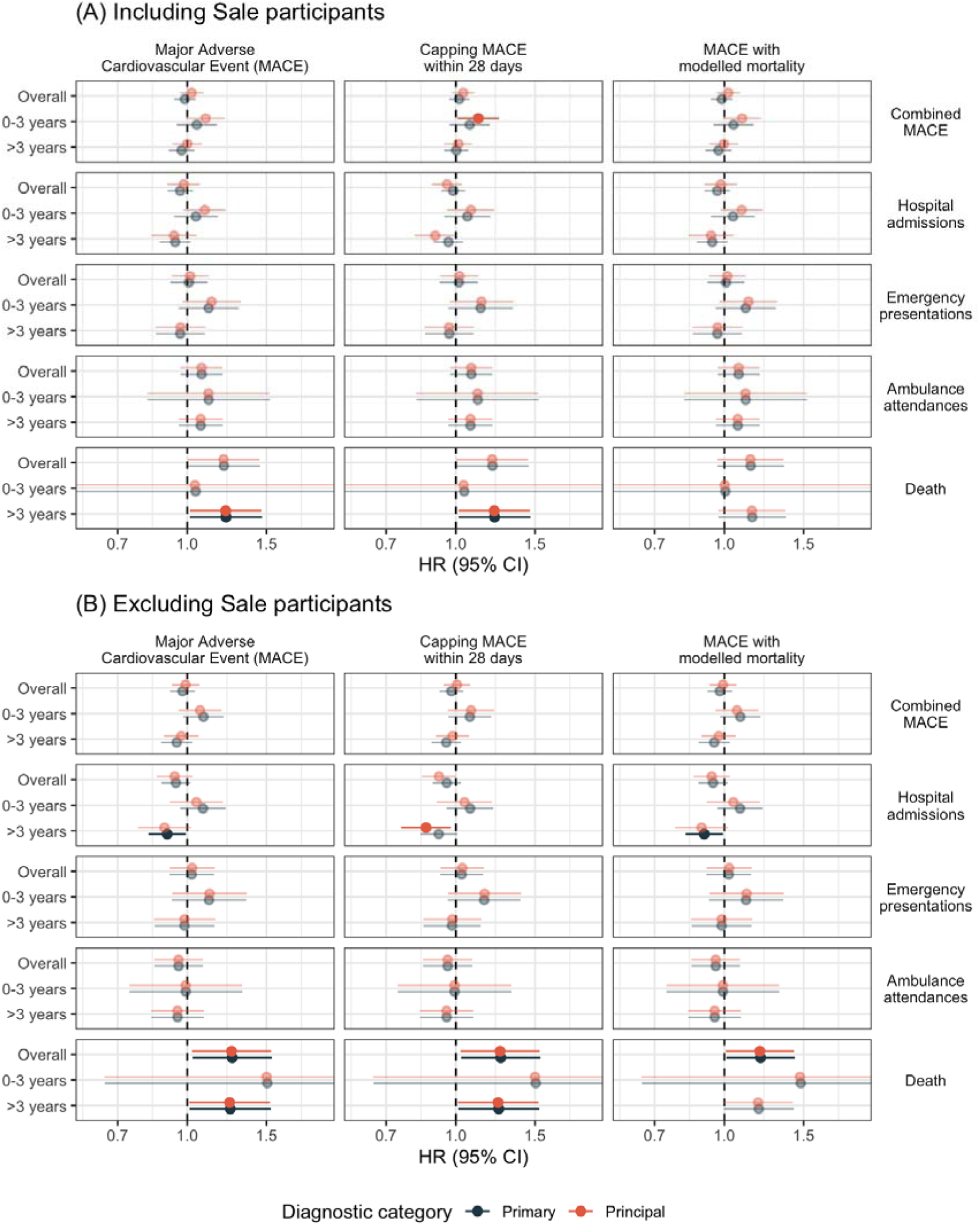
Forest plot of associations between 10µg/m^3^ increase of mine fire-related PM_2.5_ exposure and effects on MACE outcomes, for all follow-up period and stratified by time, for (a) analyses including participants from control site Sale and (b) analysis excluding participants from control site Sale. Note: bold points are statistically significant at *p* < 0.05. The first column presents the results of the MACE analysis with no modifications. The second column presents an analysis where MACE occurring within a 28-day period was counted as one event. The third column presents an analysis where MACE included modelled causes of death if they were unknown.

In contrast, for the sub-category of cardiovascular deaths, the risks were significantly elevated across the entire follow-up period (HR range: 1.14-1.26). This effect was most consistently detected in the 3 to 8 years post-fire time strata.

## Discussion

We investigated whether exposure to fire-related PM_2.5_ from the Hazelwood coal mine fire increased the incidence of MACE. There was some evidence that PM_2.5_ exposure was associated with higher MACE in the 0-3 years post-fire period, but no indication that this increase was sustained in the longer-term 3-8 years following the mine fire. However, there was evidence that PM_2.5_ exposure increased the risk of cardiovascular death in the longer-term.

These findings build on previous Hazelwood analyses, which, while not entirely consistent, generally found evidence that exposure to PM_2.5_ from the coal mine fire increased the short to medium-term risk of cardiovascular events, with limited evidence of longer-term effects.^16,19,23,43^ The findings were also consistent with another recent Hazelwood analysis, which found that the mine fire increased long-term risk of cardiovascular death,^25^ but built on them by demonstrating that the risk was elevated in the 3-8 years post-fire stratification period, meaning the harm persisted in the longer term.

Immortal time bias presents a challenge for cohort studies that recruit participants following potentially deadly disasters. We previously found evidence of premature mortality arising from PM_2.5_ exposure, particularly of cardiovascular causes, before potential participants could be recruited into the study.^43^ Counterintuitively, this phenomenon can make deadly exposures appear neutral or even protective since the surviving cohort has been depleted of its more vulnerable members; in aggregate, they artificially appear healthier than if the more vulnerable members had survived.^44^ The implications for our study include a likely underestimate in PM_2.5_ exposure effects, particularly regarding cardiovascular deaths. The relatively late recruitment into the cohort (∼2.5-3 years post-fire) also explains why confidence intervals for cardiovascular death were so wide in the first 3 years post-fire, as there would be relatively few cohort members who could have died in the short timeframe (∼0-6 months) between recruitment and stratification at 3 years.

While we only observed a sustained effect of fire-related PM_2.5_ on cardiovascular-related deaths, this did not rule out long-term effects on non-fatal MACE. Health service use is an imperfect measure of health needs, especially when there are financial constraints to accessing care. While public hospitals are free at the point of care for Australian citizens and permanent residents, there are still challenges to access in regional areas, including availability of services, costs and time required for transport.^45^ In addition, ambulance call-outs in Victoria can cost thousands of dollars for those who do not have membership, and are more expensive in regional areas.^46^ The incidence of cardiovascular conditions, even MACE, identified in this study through health service use may therefore be an underestimate and be confounded by socioeconomic differences. Morwell is one of the most socioeconomically deprived communities in Australia,^21^ and deprivation has been exacerbated by the loss of major industries following the closure of the Hazelwood coal mine and power plant in 2017.^21^

### Policy and clinical implications

The Hazelwood Health Study has yielded evidence to suggest that the mine fire increased the long-term risk of cardiovascular deaths. This should warrant a response to minimise future exposure and risk in the already exposed. Targeted health education campaigns could prepare communities for such events, focusing on strategies to reduce exposure by creating clean indoor air spaces, using air purifiers, and following early evacuation guidelines when necessary. Special emphasis should be placed on reaching individuals at greater risk of adverse cardiovascular outcomes, including those with pre-existing cardiopulmonary conditions, through community health initiatives and partnerships with healthcare providers. This group may be disproportionately affected and could benefit most from proactive measures.

Expanding access to general practitioners (GP) and primary care-based cardiovascular risk assessments, along with more accessible cardiovascular assessment and diagnostic procedures in those identified as high-risk (e.g., echocardiograms, chest CT scans, Holter monitoring, and stress tests), would enable early detection and better management of risk factors. In the aftermath of fires on the scale of the Hazelwood coal mine or larger, communities should be supported with cardiac assessments with their local GP to identify latent or emerging cardiovascular issues. Given how quickly these effects were detectable within 6 months of the fire,^24^ the best time would have been as soon as the fire was under control. However, the present analysis and another long-term study^25^ indicate there is an ongoing need to monitor cardiovascular health in communities exposed to smoke from landscape fires. Such programs could be integrated into public health responses following fire events, ensuring that those exposed receive timely and accessible care to mitigate long-term adverse outcomes.

Fire-impacted communities would benefit from improved care coordination and access to comprehensive primary healthcare services. Multidisciplinary healthy living centres could improve management of chronic conditions such as hypertension and diabetes, which are key contributors to cardiovascular risk.^47^ These centres could provide integrated services, including health assessments, referrals to specialists, dietary counselling, blood pressure monitoring and exercise programs aimed at promoting healthy lifestyles. Addressing modifiable risk factors and improving access to coordinated care could reduce the burden of cardiovascular disease over the long term, which will ultimately reduce the burden of non-communicable diseases, and the cost to the healthcare system.

## Strengths and limitations

This analysis has several strengths. Firstly, it combined data from a survey providing individual-level estimates of smoke exposure and important confounders with major administrative health databases: ambulance, emergency presentations, hospital admissions, and mortality.

The effect of immortal time bias has been discussed above in detail and remains a weakness of the analysis. However, it has a predictable biasing effect, and we could reasonably interpret this as underestimating the association between PM_2.5_ exposure and cardiovascular deaths. As a self-report survey, recall bias or general inaccuracy in time-location diaries possibly limited the accuracy of PM_2.5_ exposure estimates. There was also the possibility of diagnostic coding errors in the source health service and mortality data. Low statistical power due to a relatively modest sample size and rare outcomes also limited our ability to detect effects.

## Conclusions

Our analyses indicate a possible short-term increase in MACE-related health service use, and a sustained increase in cardiovascular deaths. This builds on our previous findings, with excess deaths reinforcing our concerns about the potential long-term cardiovascular risks of exposure to smoke from landscape fires. This emphasises the need for improved diagnostic outreach and health monitoring in populations heavily exposed to pollution and better integration of air quality data with cardiovascular health studies. Public health policies should consider how to address long-term effects of environmental disasters such as the Hazelwood coal mine fire, in terms of prevention, response and recovery. Recent landscape fire disasters, such as the one that recently devastated Los Angeles, are a stark reminder that this problem is a global concern that will not go away and likely only become more pressing.

## Supporting information

STROBE checklist

## Data Availability

HHS study data are confidential and cannot be publicly shared. Analytical code will be archived on a public repository.

https://bridges.monash.edu/articles/online_resource/Hazelwood_MACE_study_analytical_code/28629245/1

## Back matter

## Acknowledgements

Authors thank the Latrobe Valley and Gippsland communities for their support and participation in the Hazelwood Health Study. Recruitment into the Adult Survey was overseen by Susan Denny.

## Availability of data and material

HHS study data are confidential and cannot be publicly shared. Analytical code will be archived on a public repository.^48^

## Supplementary materials

**Table S1.**
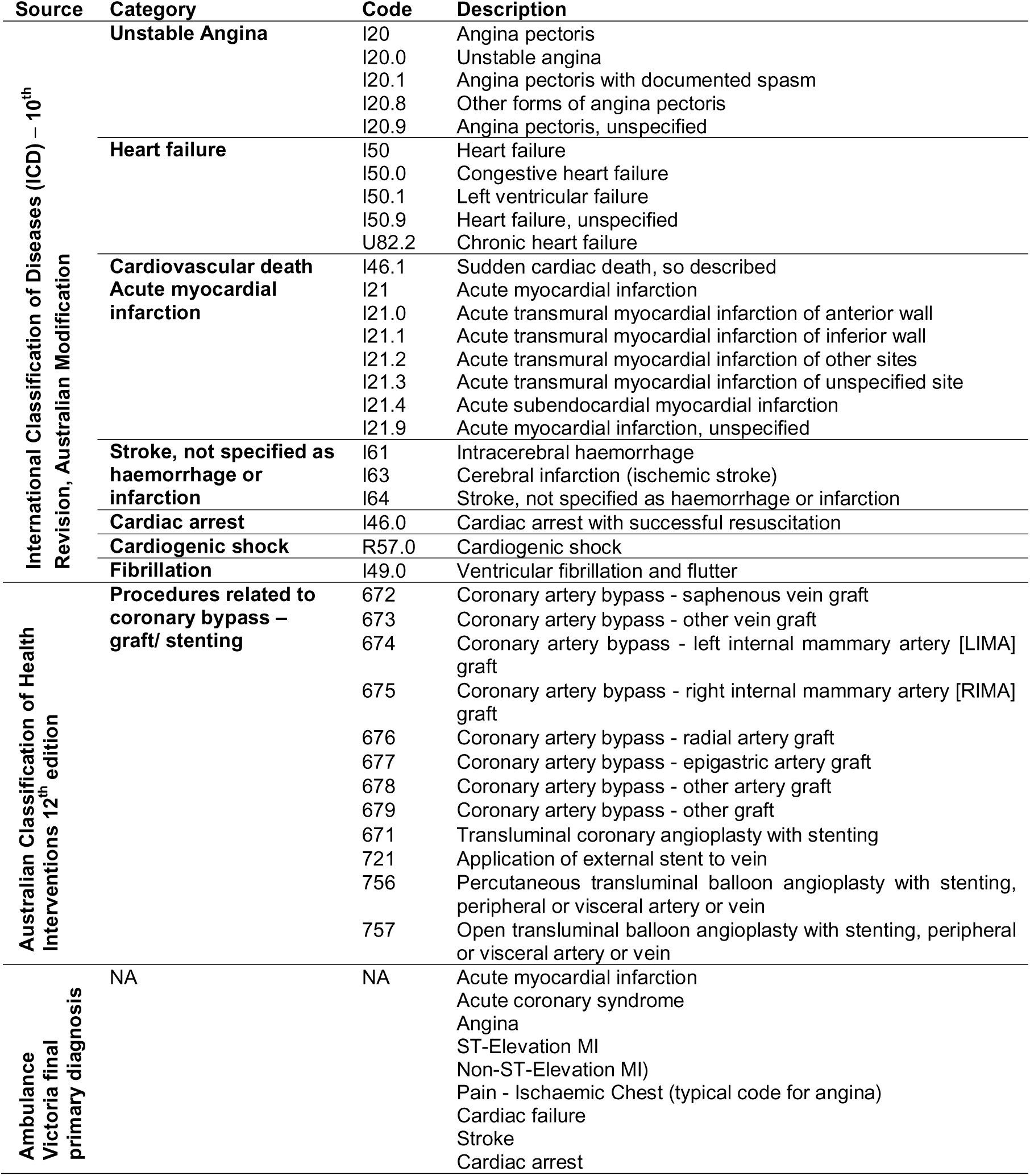
List of diagnostic and procedure codes to identify major adverse cardiovascular events (MACE)

**Table S2.**
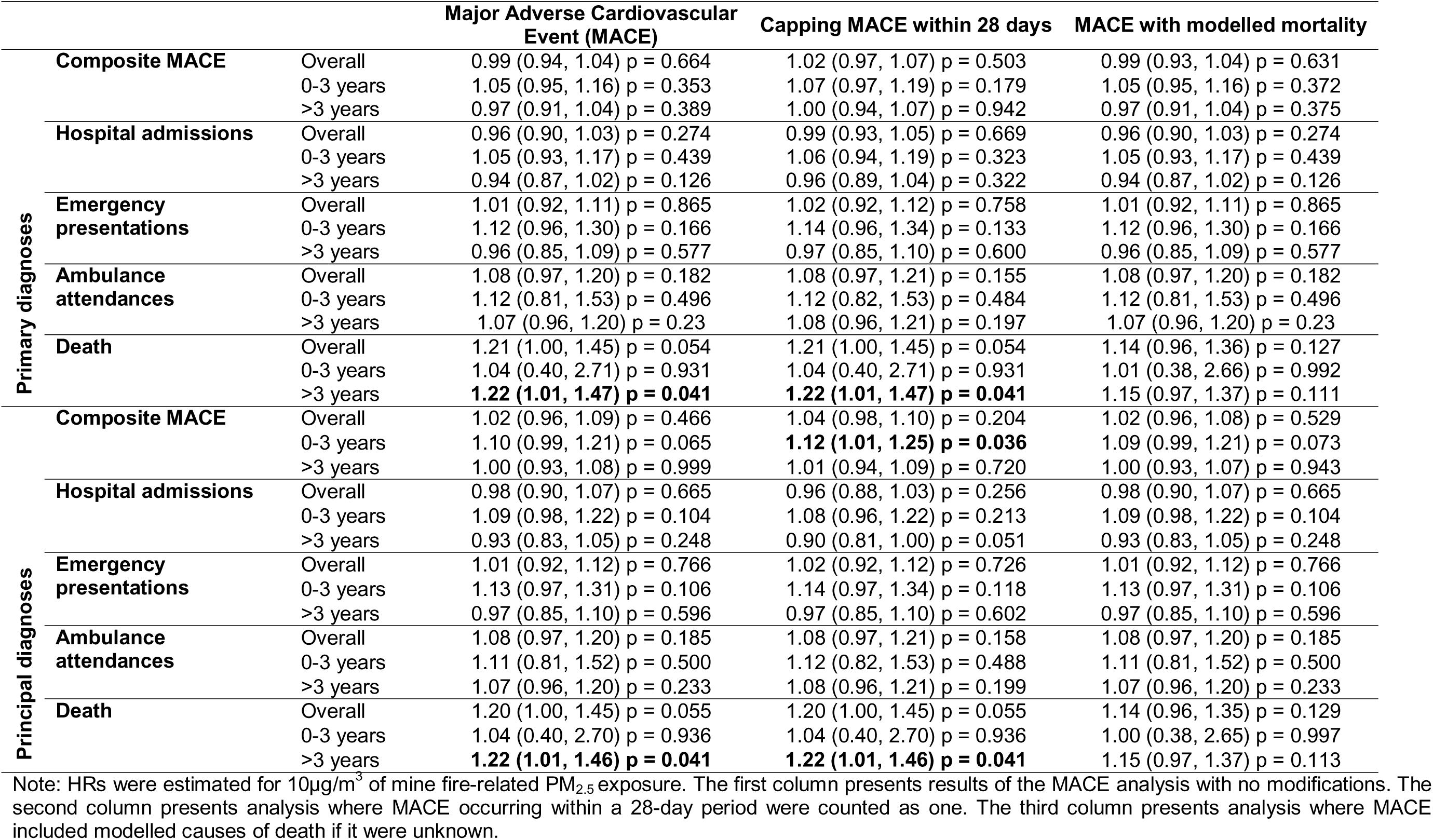
Survival analysis models for individual health service data - underlying data for Figure 1 (A)

**Table S3.**
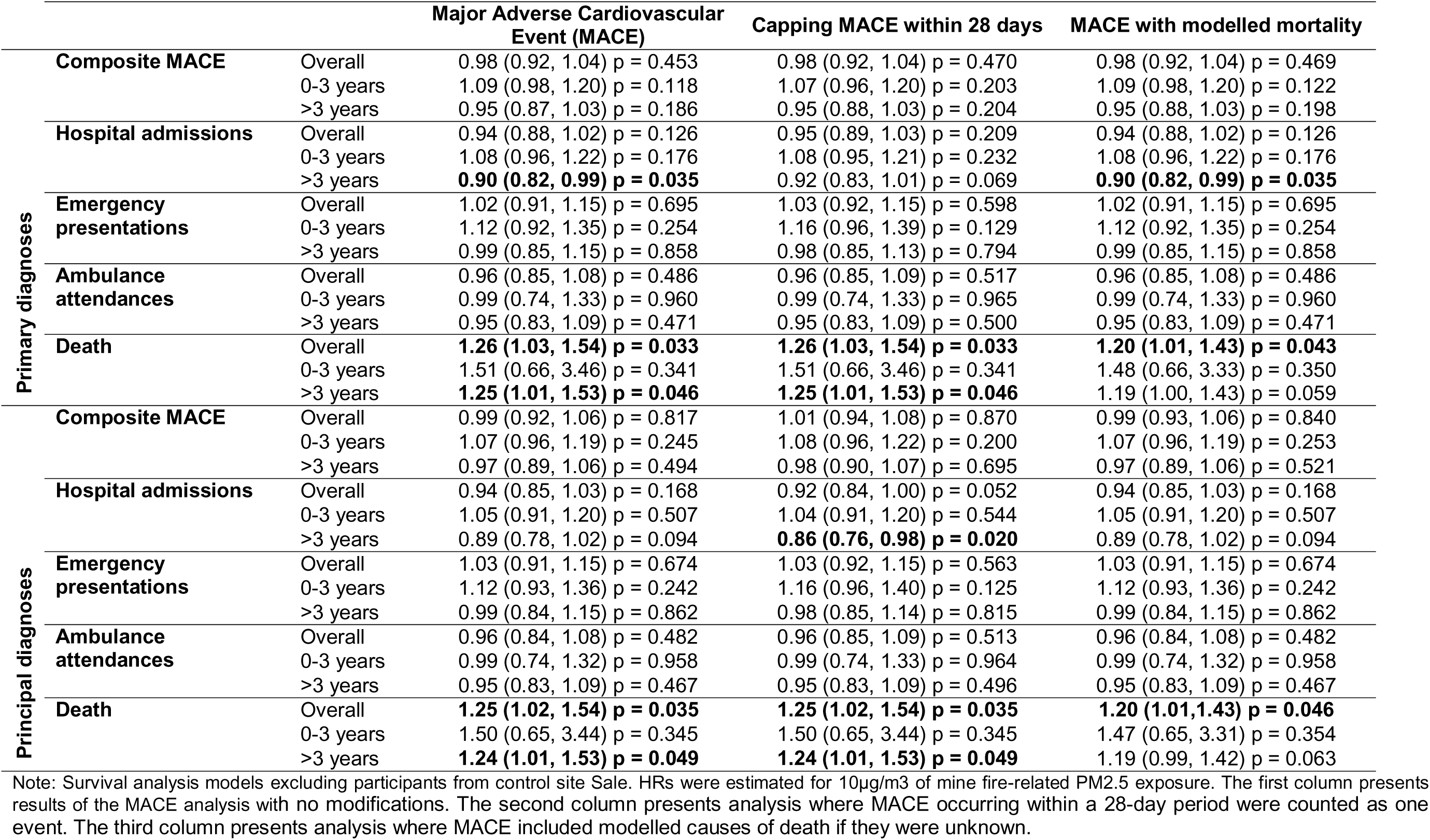
Survival analysis models for individual health service data - underlying data for Figure 1 (B)

**Table S4.**
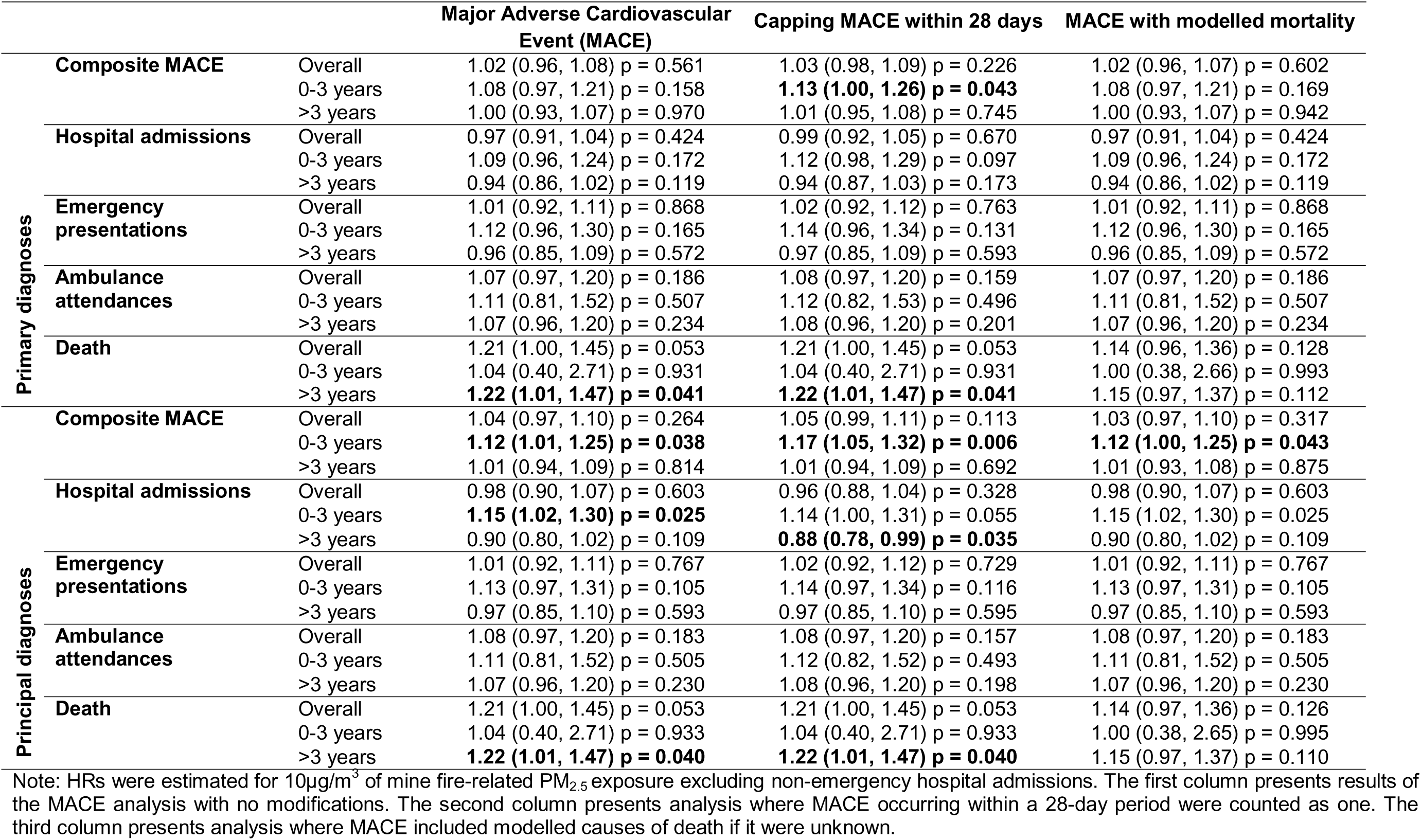
Survival analysis models for individual health service data - underlying data for Figure S1 (A)

**Table S5.**
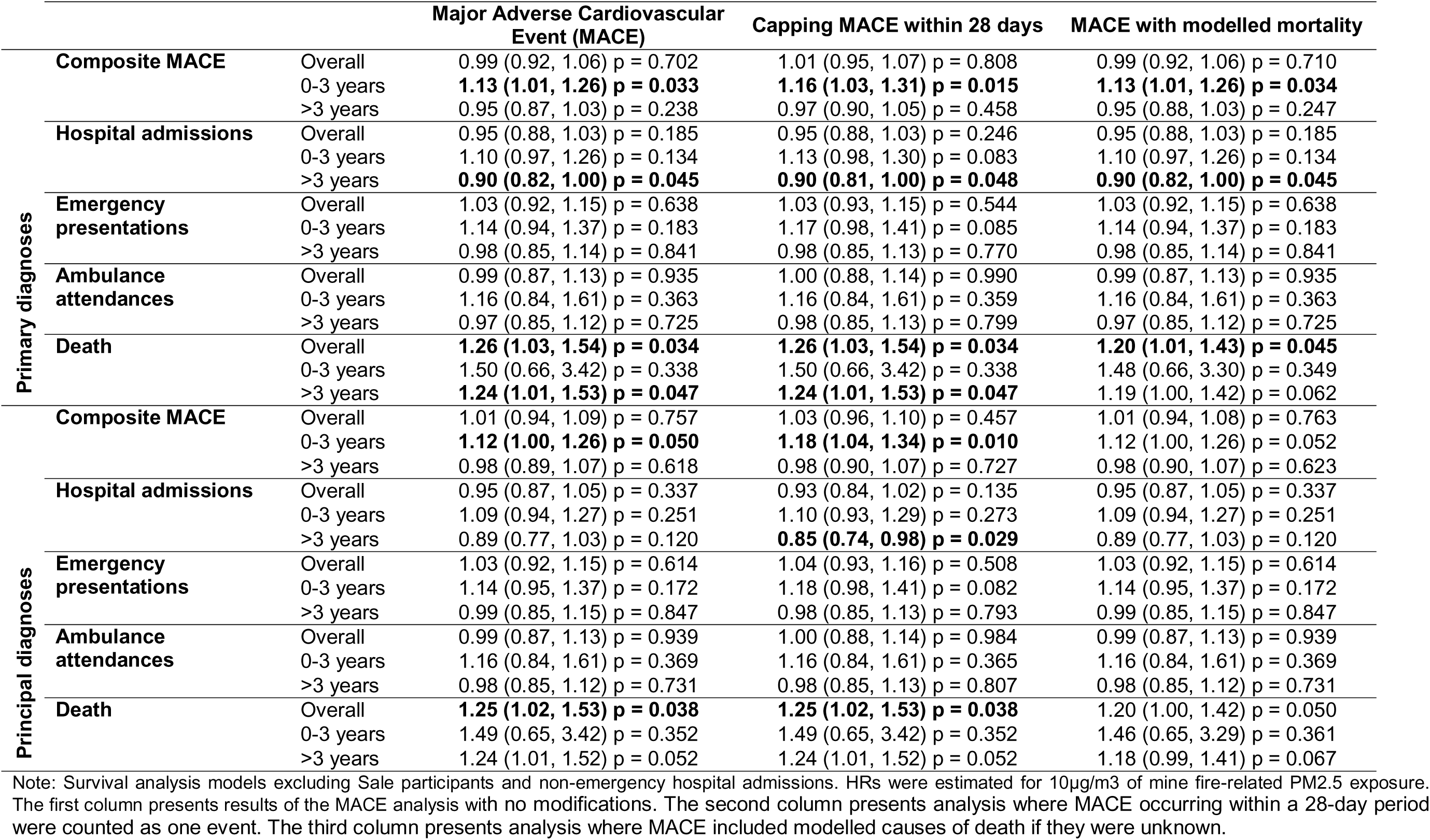
Survival analysis models for individual health service data - underlying data for Figure S1 (B)

**Figure S2.**
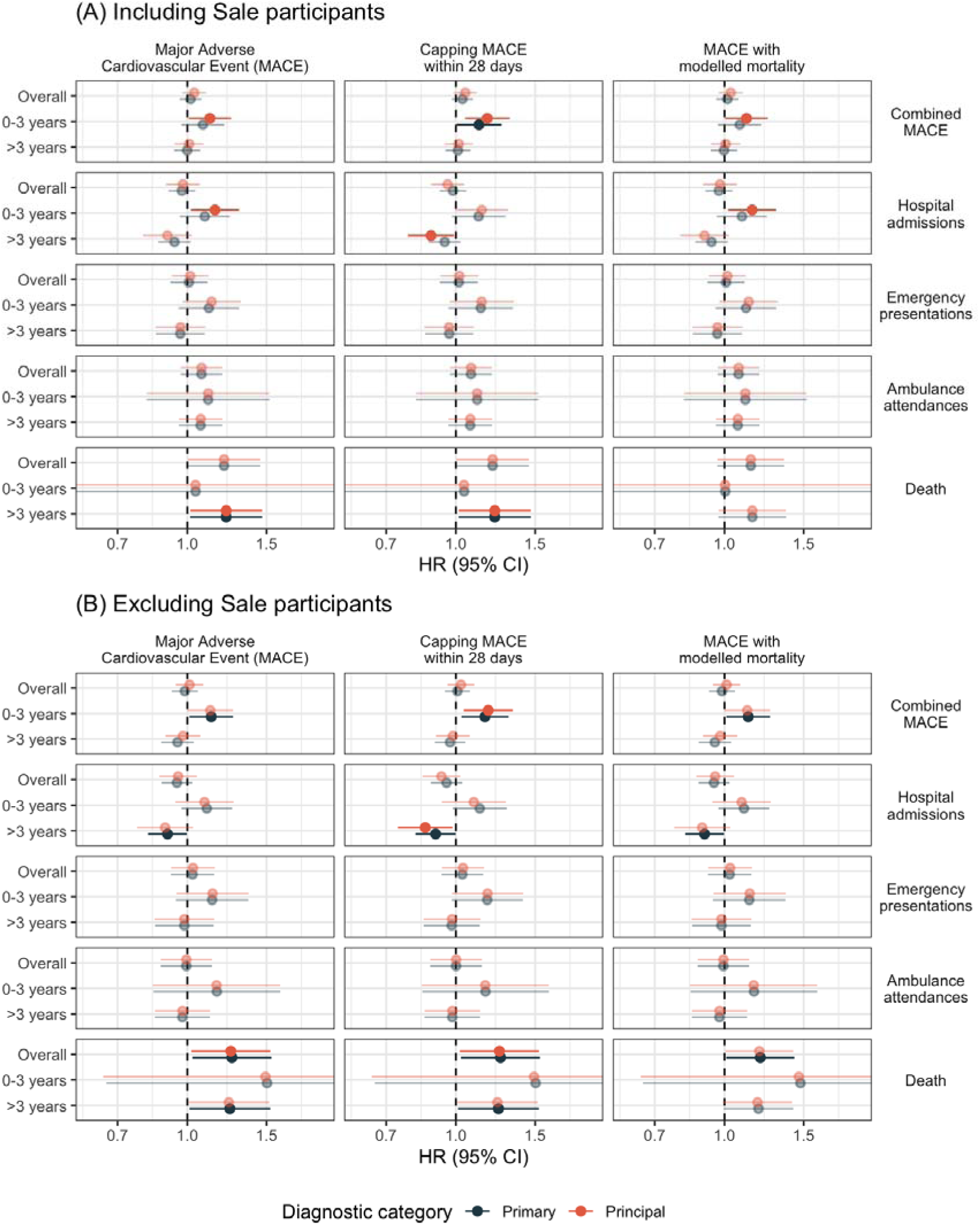
Forest plot of associations between 10µg/m^3^ of PM_2.5_ exposure and effect on MACE outcomes including predicted cardiovascular-related death, sensitivity analysis excluding non-emergency hospital admissions, for (a) analyses including participants from control site Sale and (b) analysis excluding participants from control site Sale. Note: bold points are statistically significant at *p* < 0.05. The first column presents results of the MACE analysis with no modifications. The second column presents analysis where MACE occurring within a 28-day period were counted as one event. The third column presents analysis where MACE included modelled causes of death if they were unknown.

