## Supplementary material for "Effects of PM_2.5_ from coal mine fire smoke on long-term incidence of major adverse cardiovascular events (MACE)": STROBE checklist

dSTROBE Statement—checklist of items that should be included in reports of observational studies

|  | Item No. | Recommendation | Page  No. | Relevant text from manuscript |
| --- | --- | --- | --- | --- |
| **Title and abstract** | 1 | (*a*) Indicate the study’s design with a commonly used term in the title or the abstract | 2 | “In this cohort study…” |
|  |  | (*b*) Provide in the abstract an informative and balanced summary of what was done and what was found | 2 | This is described in both the Methods and Results subsections of the abstract. |
| Introduction | | | |  |
| Background/rationale | 2 | Explain the scientific background and rationale for the investigation being reported | 3-4 | “…studies of effects on cardiovascular health services covering medications, hospital admissions, and ambulance attendances, have been mixed, indicating both an increase in use and no effect.^14–19^” |
| Objectives | 3 | State specific objectives, including any prespecified hypotheses | 4 | “In this analysis, we focused on major adverse cardiovascular events (MACE), one of the most common primary endpoints in cardiovascular research.^26^” |
| Methods | | | |  |
| Study design | 4 | Present key elements of study design early in the paper | 5 | “…we used the Victorian Electoral Roll to identify and recruit 4056 adult residents of areas near the Hazelwood coal mine fire into a cohort known as the Adult Survey.” |
| Setting | 5 | Describe the setting, locations, and relevant dates, including periods of recruitment, exposure, follow-up, and data collection | 5 | “All four data sources were linked to Adult Survey participants covering the period from January 1, 2009, to June 30, 2022, with the follow-up period defined as the interval between March 31, 2014 (end of the mine fire) and June 30, 2022. Due to differences in healthcare accessibility and long-term changes in socioeconomic status after the mine fire between Morwell and Sale, the main analysis focused on 2115 cohort members from Morwell.” |
| Participants | 6 | (*a*) *Cohort study*—Give the eligibility criteria, and the sources and methods of selection of participants. Describe methods of follow-up | 5 | “This study relied on data from the Hazelwood Health Study Adult Survey, which has been described previously.^29,30^” |
|  |  | (*b*) *Cohort study*—For matched studies, give matching criteria and number of exposed and unexposed | 5 | “This included 3096 residents of Morwell, the primary exposure site, and 960 residents of Sale, an unexposed town around 60km to the northeast. Of these, 2725 (2115 from Morwell, 610 from Sale) agreed to have their data linked to a range of administrative health datasets.” |
| Variables | 7 | Clearly define all outcomes, exposures, predictors, potential confounders, and effect modifiers. Give diagnostic criteria, if applicable | 5-7 | Under subheadings “Exposure to mine fire-related PM_2.5_”, “Outcomes”, and “Confounders”. |
| Data sources/ measurement | 8* | For each variable of interest, give sources of data and details of methods of assessment (measurement). Describe comparability of assessment methods if there is more than one group | 5-7 | Under subheadings “Exposure to mine fire-related PM_2.5_”, “Outcomes”, and “Confounders”. |
| Bias | 9 | Describe any efforts to address potential sources of bias | 7-8 | Under subheadings “Confounders”, “Analysis” |
| Study size | 10 | Explain how the study size was arrived at | 5 | “In summary, we used the Victorian Electoral Roll to identify and recruit 4056 adult residents of areas near the Hazelwood coal mine fire into a cohort known as the Adult Survey. This included 3096 residents of Morwell, the primary exposure site, and 960 residents of Sale, an unexposed town around 60km to the northeast. Of these, 2725 (2115 from Morwell, 610 from Sale) agreed to have their data linked to a range of administrative health datasets.” |

| Quantitative variables | 11 | Explain how quantitative variables were handled in the analyses. If applicable, describe which groupings were chosen and why | 5-8 | Under subheadings “Exposure to mine fire-related PM_2.5_”, “Outcomes”, “Confounders”, and “Analysis”. |
| --- | --- | --- | --- | --- |
| Statistical methods | 12 | (*a*) Describe all statistical methods, including those used to control for confounding | 7-8 | Under subheading “Analysis” |
|  |  | (*b*) Describe any methods used to examine subgroups and interactions | NA |  |
|  |  | (*c*) Explain how missing data were addressed | 8 | “Missing data were addressed using Multivariate Imputation by Chained Equations (MICE), producing 20 imputed datasets that were analysed separately and had their results pooled using Rubin's rules.^39^ All analyses were conducted using R in RStudio.^40,41^” |
|  |  | (*d*) *Cohort study*—If applicable, explain how loss to follow-up was addressed | NA | Data linkage that did not require follow-up. |
|  |  | (*e*) Describe any sensitivity analyses | 8 | “We also analysed each approach to the MACE definition separately (based on primary versus principal diagnosis; capping events within a rolling 28 days; with predicted cardiovascular-related mortality). Analysis of MACE excluding non-emergency hospital procedures served as a sensitivity analysis i.e., restricted to ambulance, ED, death and emergency hospitalisations.” |
| Results | | | | |
| Participants | 13* | (a) Report numbers of individuals at each stage of study—eg numbers potentially eligible, examined for eligibility, confirmed eligible, included in the study, completing follow-up, and analysed | 9-10 | Table 1 |
|  |  | (b) Give reasons for non-participation at each stage | 6 | “In summary, we used the Victorian Electoral Roll to identify and recruit 4056 adult residents of areas near the Hazelwood coal mine fire into a cohort known as the Adult Survey. This included 3096 residents of Morwell, the primary exposure site, and 960 residents of Sale, an unexposed town around 60km to the northeast. Of these, 2725 (2115 from Morwell, 610 from Sale) agreed to have their data linked to a range of administrative health datasets.” |
|  |  | (c) Consider use of a flow diagram | NA |  |
| Descriptive data | 14* | (a) Give characteristics of study participants (eg demographic, clinical, social) and information on exposures and potential confounders | 9-10 | Table 1 |
|  |  | (b) Indicate number of participants with missing data for each variable of interest | 9-10 | Table 1 |
|  |  | (c) *Cohort study*—Summarise follow-up time (eg, average and total amount) | 5 | “…the follow-up period defined as the interval between March 31, 2014 (end of the mine fire) and June 30, 2022.” |
| Outcome data | 15* | *Cohort study*—Report numbers of outcome events or summary measures over time | 9-10 | Table 1 |
|  |  | *Case-control study—*Report numbers in each exposure category, or summary measures of exposure | *NA* |  |
|  |  | *Cross-sectional study—*Report numbers of outcome events or summary measures | *NA* |  |
| Main results | 16 | (*a*) Give unadjusted estimates and, if applicable, confounder-adjusted estimates and their precision (eg, 95% confidence interval). Make clear which confounders were adjusted for and why they were included | 12; Supp materials | Figure 1; Table S3 |
|  |  | (*b*) Report category boundaries when continuous variables were categorized | 9-10 | Table 1 |
|  |  | (*c*) If relevant, consider translating estimates of relative risk into absolute risk for a meaningful time period | NA |  |

Continued on next page

| Other analyses | 17 | Report other analyses done—eg analyses of subgroups and interactions, and sensitivity analyses | 11; Suppl materials | “Sensitivity analyses produced similar results. These are illustrated in Figure S1.”; Table S1 |
| --- | --- | --- | --- | --- |
| Discussion | | | | |
| Key results | 18 | Summarise key results with reference to study objectives | 13 | “There was some evidence that PM_2.5_ exposure was associated with higher MACE in the 0-3 years post-fire period, but no indication that this increase was sustained in the longer term 3-8 years following the mine fire. An important exception was cardiovascular deaths, which were elevated across the entire follow-up period.” |
| Limitations | 19 | Discuss limitations of the study, taking into account sources of potential bias or imprecision. Discuss both direction and magnitude of any potential bias | 13; 15 | “In our study, there was evidence of premature mortality arising from PM_2.5_ exposure, particularly of cardiovascular causes, before potential participants could be recruited into the study.^43^ Counterintuitively, this phenomenon can make deadly exposures appear neutral or even protective since the surviving cohort is derived from a healthier population.^44^ The implications for our study include a likely underestimate in PM_2.5_ exposure effects, particularly regarding cardiovascular deaths.”; “Strengths and limitations” subsection |
| Interpretation | 20 | Give a cautious overall interpretation of results considering objectives, limitations, multiplicity of analyses, results from similar studies, and other relevant evidence | 16 | “Our analyses indicate a possible short-term increase in MACE-related health service use, and a sustained increase in cardiovascular deaths.” |
| Generalisability | 21 | Discuss the generalisability (external validity) of the study results | 13 | “The implications for our study include a likely underestimate in PM_2.5_ exposure effects, particularly regarding cardiovascular deaths.” |
| Other information | |  | | |
| Funding | 22 | Give the source of funding and the role of the funders for the present study and, if applicable, for the original study on which the present article is based | 17 | “This work was funded by the Victorian Department of Health. The paper presents the views of the authors and does not represent the views of the Department.” |

*Give information separately for cases and controls in case-control studies and, if applicable, for exposed and unexposed groups in cohort and cross-sectional studies.

**Note:** An Explanation and Elaboration article discusses each checklist item and gives methodological background and published examples of transparent reporting. The STROBE checklist is best used in conjunction with this article (freely available on the Web sites of PLoS Medicine at http://www.plosmedicine.org/, Annals of Internal Medicine at http://www.annals.org/, and Epidemiology at http://www.epidem.com/). Information on the STROBE Initiative is available at www.strobe-statement.org.
